# Effect of xenon and argon inhalation on erythropoiesis and steroidogenesis: a systematic review

**DOI:** 10.1101/2022.03.30.22273135

**Authors:** Eduard Bezuglov, Ryland Morgans, Ruslan Khalikov, Vladislav Bertholz, Anton Emanov, Oleg Talibov, Evgeniy Astakhov, Artemii Lazarev, Maria Shoshorina

## Abstract

**Background:** Xenon and argon inhalation were included in the WADA Prohibited List in 2014 due to the reported positive effects on erythropoiesis and steroidogenesis occurs as a result of the use of these substances. Currently, xenon is on the WADA Prohibited List notable affecting erythropoiesis as a Hypoxia-inducible factor (HIF) activating agent. At the same time, argon inhalation was allowed again in 2019. Thus, the systematic review of studies supporting these notions is of interest.

**Methods:** A thorough search for articles on the effects of xenon and argon inhalation on erythropoiesis and steroidogenesis, as well as their negative effects on human health and methods of their detection in body fluids was conducted. Pubmed, Google Scholar databases and the Cochrane Library were researched, as well as the special research section of the WADA website. The search was conducted in accordance with PRISMA guidelines. All articles in English published between 2000 and 2021 were analyzed, as well as reference studies meeting the search criteria.

**Results:** At present, there are only two publications in healthy human subjects evaluating the effects of xenon inhalation on erythropoiesis were found with no conclusive evidence of a positive effect on erythropoiesis. Both articles were published after 2014 when the gases were included on the WADA Prohibited List. Both articles had a high risk of bias. There were no studies on the effect of argon inhalation on erythropoiesis. No studies were found on the effect of xenon or argon inhalation on steroidogenesis in healthy subjects. No studies related to the effects of xenon or argon inhalation on erythropoiesis and steroidogenesis were found on the WADA website.

**Conclusion:** There is still inconclusive evidence to support the administration of xenon and argon inhalations on erythropoiesis and steroidogenesis and their positive effects on health. Further research is needed to establish the effects of these gases. Additionally, improved communication between the anti-doping authorities and all key stakeholders is required to support the inclusion of various substances on the Prohibited List.

## Introduction

Xenon is an inert gas that is present in the atmosphere. There are nine xenon isotopes, the most common of which is Xe 132. The narcotic effect of xenon has been previously reported in rats by inhaling 67% xenon and 33% oxygen [27]. The narcotic properties of the xenon-oxygen mixture (78% xenon 22% oxygen) were also previously described in experiments on mice [26]. The first studies on humans showed that inhalation of a mixture of 80% xenon and 20% oxygen resulted in full anesthesia in 3-5 minutes [14].

In Russia, xenon has been officially approved for use as an inhalation anesthetic since 1999. By 2012, more than 10,000 surgical operations have been performed with the use of xenon anesthesia [12]. Xenon does not have teratogenic, mutagenic, carcinogenic, or allergenic properties and does not affect respiratory function [53],[31],[13],[22]. The official recognition of xenon as an inhalation anesthetic and subsequent clinical use of xenon has revealed several physiological effects, which has contributed to its fairly widespread introduction into clinical practice. There have been numerous potential applications of xenon including: treatment of withdrawal symptoms, drug addiction and depression, chemotherapy, analgesia and their possible cardiac, renal, and neuroprotective properties in the elderly population [15],[47],[10],[38],[21],[3],[32]. In addition to its use in anesthesiology, in Russia, it has been actively studied as an adaptive agent for recovery from extreme physical exertion in athletes and military personnel [33],[30].

In September 2014, inhalation of xenon and another inert gas, argon with oxygen (hereinafter referred to as IX and IA) was added to the WADA Prohibited List under section S2.1 as hypoxia-inducible factor (HIF) stabilizers and activators [49]. To fully establish the circumstances that led to the inclusion of IX and IA to the Prohibited List, a clear chronology of events occurring in 2014 must be established. On the 7th February 2014, the Russian city of Sochi opened the Winter Olympics and concurrently an article appeared in the “Economist” stating that xenon is being employed as a way to improve athletic performance and that Russian athletes participating in the Sochi Olympics could use xenon inhalation as a performance-enhancing substance [11]. The article claimed that the official document released in 2010 by the State Research Institute of the Ministry of Defense set out guidelines for the administration of the gas to athletes. In the fore-mentioned report (the link to which was not provided) it was allegedly recommended to use xenon before competition to eliminate lethargy and sleep disorders and to improve physical recovery. It was recommended that xenon and oxygen should be used in a 50:50 ratio when inhaled for a few minutes before going to sleep. The report also stated that the effect of the gas lasts for 48-72 hours, so it was recommended to repeat this procedure every 2-3 days.

During the Olympics, German broadcaster WDR claimed members of the Russian team at Sochi had inhaled the gas and alleged that top Russian athletes have been using xenon to improve performances since the Athens Olympics in 2004. This report, like the Economist’s earlier publication, offered no empirical evidence to support the claim that xenon had a positive effect on erythropoiesis. Furthermore, on February 27th, immediately post-Olympics, the website www.insidethegames.biz, with reference to WDR, published information that Russian athletes had used xenon during competition and that WADA President Sir Craig Reedie has said “ it will take on the issue” [1]. WADA started a process to verify these claims.

It should be emphasized that IX had never previously been a banned substance and had been openly used by Russian athletes at major international competitions and this was well-known by some WADA executives. For example, this was stated by David Howman, Director General of the World Anti-Doping Agency, who said that xenon use had been known for “years and years, before Athens 2004” but had not been previously looked at because “it wasn’t an issue that needed to be addressed” [2]. In other words, there were no questions about the use of IX and IA for many years. However, former WADA president, a member of its Foundation Board, and a member of the International Olympic Committee, Dick Pound, stated: ‘in no doubt that it is doping” [2]. Simultaneously, WADA president Sir Craig Reedy promised that “the topic of gas will already be addressed at the next meeting after the Olympics” [18]. In April 2014, following the WADA Prohibited List Committee meeting, IX and IA were recommended for inclusion on the Prohibited List, where they have been included as Hypoxia-Inducible Factor (HIF) activators since 1 September 2014 [48]. It is important to recall that they were included in the same section S2 of the Prohibited List as the long-established commonly used drugs with well-documented positive effects on erythropoiesis: erythropoietin (EPO) and darbepoetin (dEPO). Thus, IX and IA became prohibited substances in sports and a minimum two-year ban was imposed for their use.

Noteworthy, to be included on the Prohibited List, a substance or method must satisfy at least two of three criteria:

1. It has the potential to enhance or enhances sport performance;
2. It represents an actual or potential health risk to the athlete;
3. It violates the spirit of sport [52].

The third criterion is relatively vague and virtually any substance or method may qualify for it. However, at least one other criterion must be met. In this regard, the inclusion of IX and IA to the Prohibited List would have been based on WADA experts having compelling data on the positive effects on physical performance or their negative effects on human health. This could have been confirmed by WADA President Sir Craig Reedy’s comment in an interview to the Telegraph on 18 May 2014 that the ban on IX and IA was based on research available to the agency indicating their positive effects on erythropoiesis and steroidogenesis [34]. Therefore, the question arises why these data were not mentioned previously and why the decision to ban IX and IA was taken so abruptly?

The reason for the ban may have been the detection of a significant increase in its use or widespread use among athletes of any group, which may have been indicative of the unpublished evidence of its effectiveness in relation to performance or post-exercise recovery. For example, the banning of meldonium was implemented after its use was found to be extremely widespread at the 2015 European Games in Baku [43]. Therefore, one of the aims of this review was to examine the chronology of the emergence of xenon and argon detection methods in biological fluids.

The aim of this systematic review was to further examine the evidence base supporting the inclusion of the IX and IA on the Prohibited List.

## Materials and methods

Two independent expert researchers conducted the search utilising the Pubmed, Cochrane Library and Google Scholar databases. Additionally, the WADA website reporting on the effects of xenon and argon inhalation on various aspects of physical performance, erythropoiesis and steroidogenesis, and their negative effects on human health and methods for their detection in body fluids was also searched. These searches were conducted according to PRISMA guidelines. All English published articles between 2000 and 2021 were analyzed and reference articles meeting the search criteria. A citation list of all articles that met the search inclusion criteria including articles on xenon detection in body fluids in doping control was also examined.

The following words and word combinations were used for the search: “Xenon inhalation”, “Argon inhalation”, “Xenon and erythropoiesis”, “Argon and erythropoiesis”, “Xenon and steroidogenesis”, “Argon and steroidogenesis”, “Xenon and testosterone”, “Argon and testosterone”, “Xenon and erythropoietin”, “Argon and erythropotin”, “Xenon and endurance”, “Argon and endurance”, “Xenon and performance”, “Argon and performance”, “Xenon and Sport”, “Argon and Sport”, “Xenon and Athletes”, “Argon and Athletes”, “Xenon Negative Effects”, “Argon Negative Effects”, “Xenon Inhalation Side Effects”, “Xenon Inhalation Adverse Effects”, Xenon Inhalation Adverse Events” “Argon Inhalation Side Effects”, “Xenon and Detection Methods”, “Argon and Detection Methods”, “Xenon and Doping”, “Argon and Doping*”*.

The use of IX and IA in 2014 was documented in connection with Russian athletes, thus the above keywords and their combinations were also searched in the Russian-language scientific databases and CyberLeninka.

The inclusion criteria were:

1. The article is a clinical study;
2. The subjects of the study were healthy individuals;
3. The study was about the influence of xenon and argon on erythropoiesis and steroidogenesis;
4. The intervention was xenon or argon versus placebo inhalation.

Information on all disqualified athletes was also searched on the rusada.ru and major winter sports federations’ websites in which Russian athletes participated at the Sochi Olympics. This was completed to identify athletes who had been disqualified for using IX and IA since the second half of 2014, when it was banned. This time-line was adopted due to samples taken during major international competitions (including the World Winter Sports Championships and the Winter Olympics) are now retrospectively analyzed within 10 years of their conclusion. This was completed to examine the use of prohibited substances and methods not found in the original analysis following the emergence of new detection protocols.

This study was approved by the local ethical committee of the Sechenov First Moscow State Medical University (N 11-19).

All identified studies have been assessed for risk of bias using the Revised Cochrane risk-of-bias tool for randomized trials (RoB 2) [54]. Cases of disagreement in assessments of the risk of bias between the reviewers were resolved by discussion or with consultation with a third reviewer if needed.

## Results

Based on keywords, and combinations thereof, 4,979 articles were retrieved (See Figure 1). In the first phase, 19 duplicates were excluded, followed by three more articles, one of which was written in French [44] and the other two were a commentary and response to a commentary on the study by Stoppe et al. [4],[42]. Next, 4,955 articles were screened by title and abstract. Due to the inappropriateness of the topic of this study, 4,938 publications were excluded.

**Figure 1.**
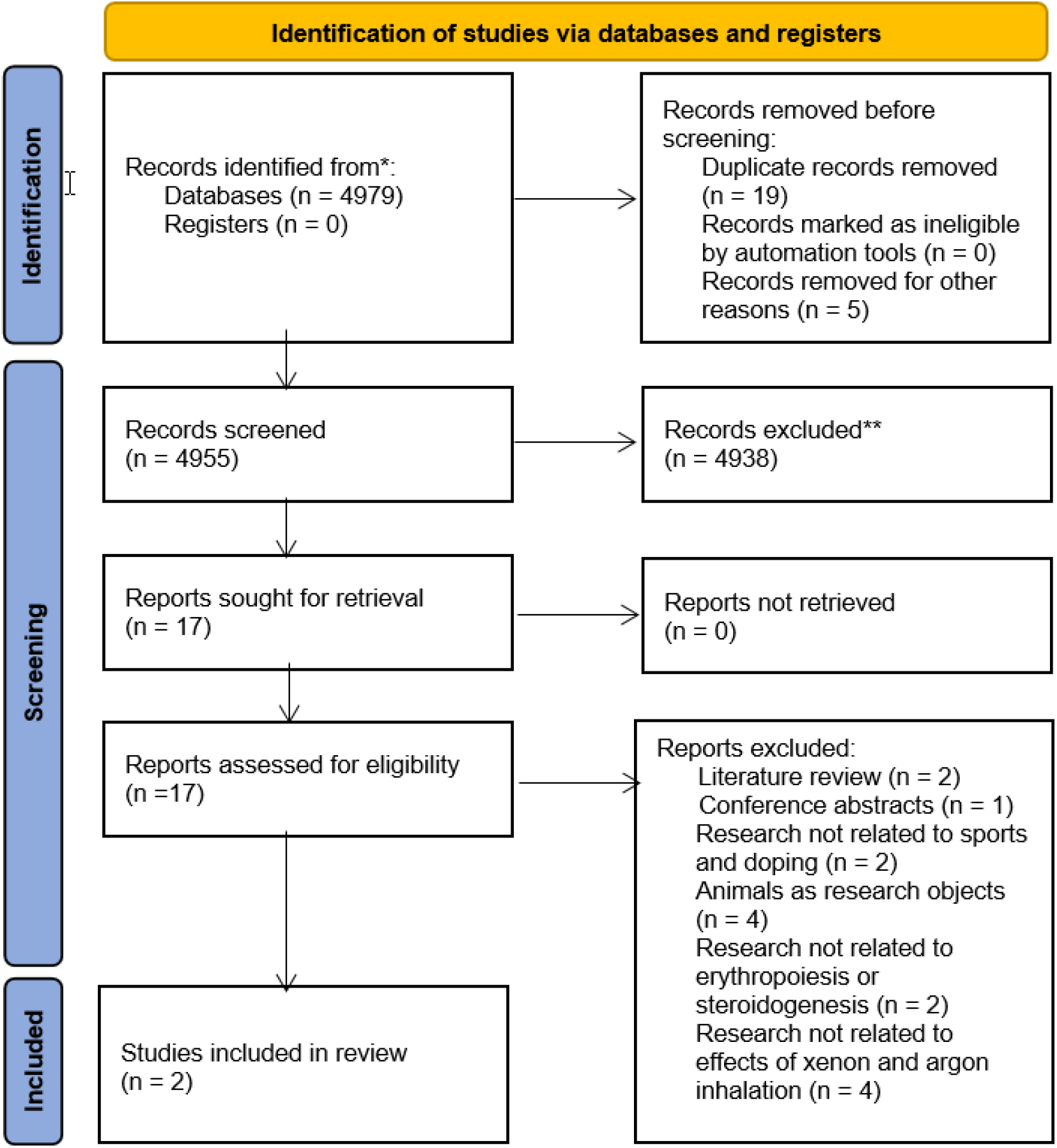
Study selection process.

Seventeen publications were then screened in detail to ensure that the following criteria were met: two studies were literature reviews [45],[19] and one study was the primary material [40] for the secondary analysis [39], which will be mentioned later in this article. Four studies on the effect of IX on erythropoiesis were performed with animals (rats and mice) [20],[29],[28] and another involving people after undergoing heart surgery [39]. In summary, only two publications were found on studies involving healthy subjects in English that evaluated the effects of xenon inhalation on erythropoiesis (See Table 1) [41],[16]. Regarding methods for detection of IX in biological fluids, several studies were found notably one sponsored by WADA [46],[35],[17],[25],[36],[24].

**Table 1.**
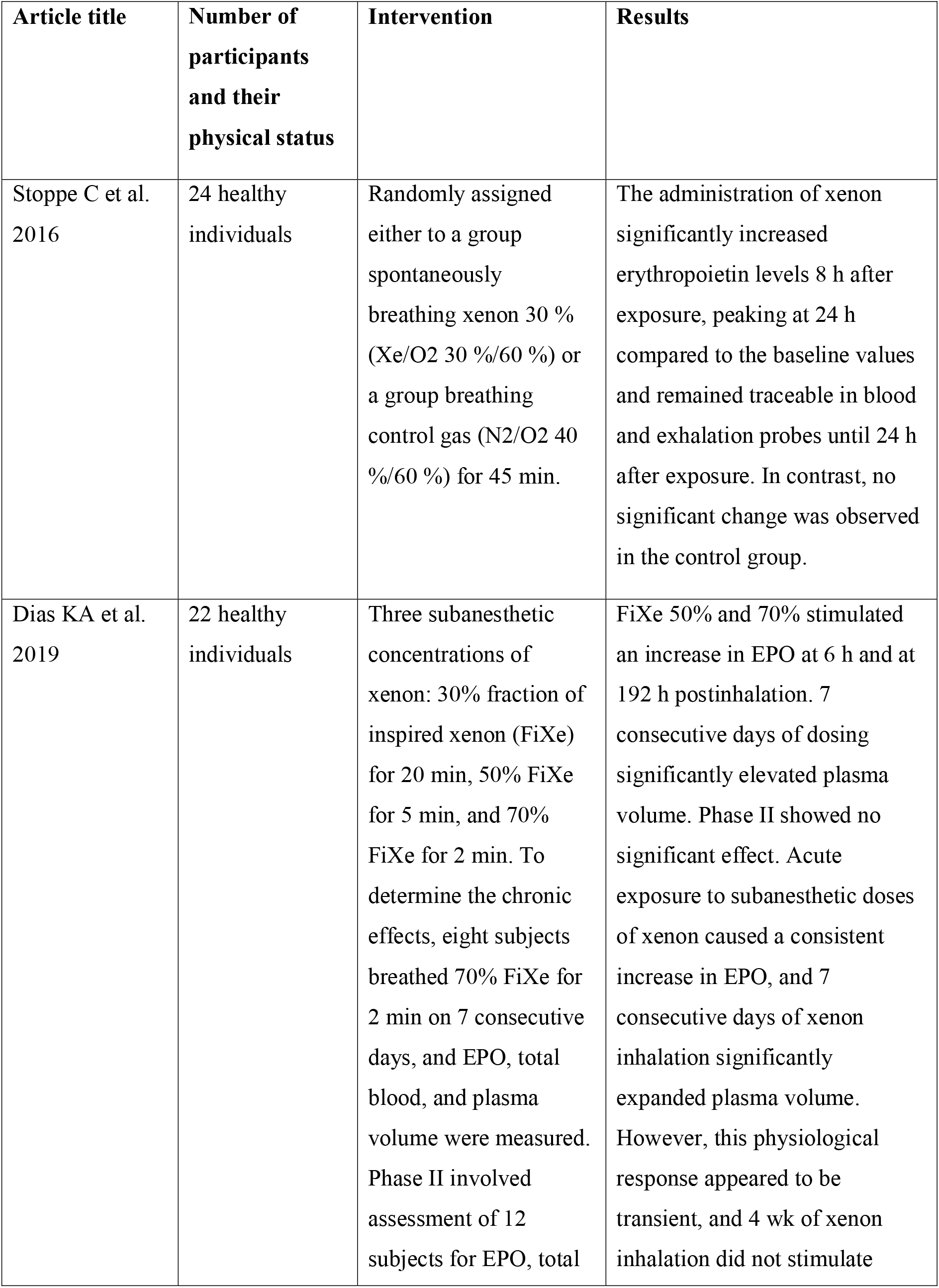
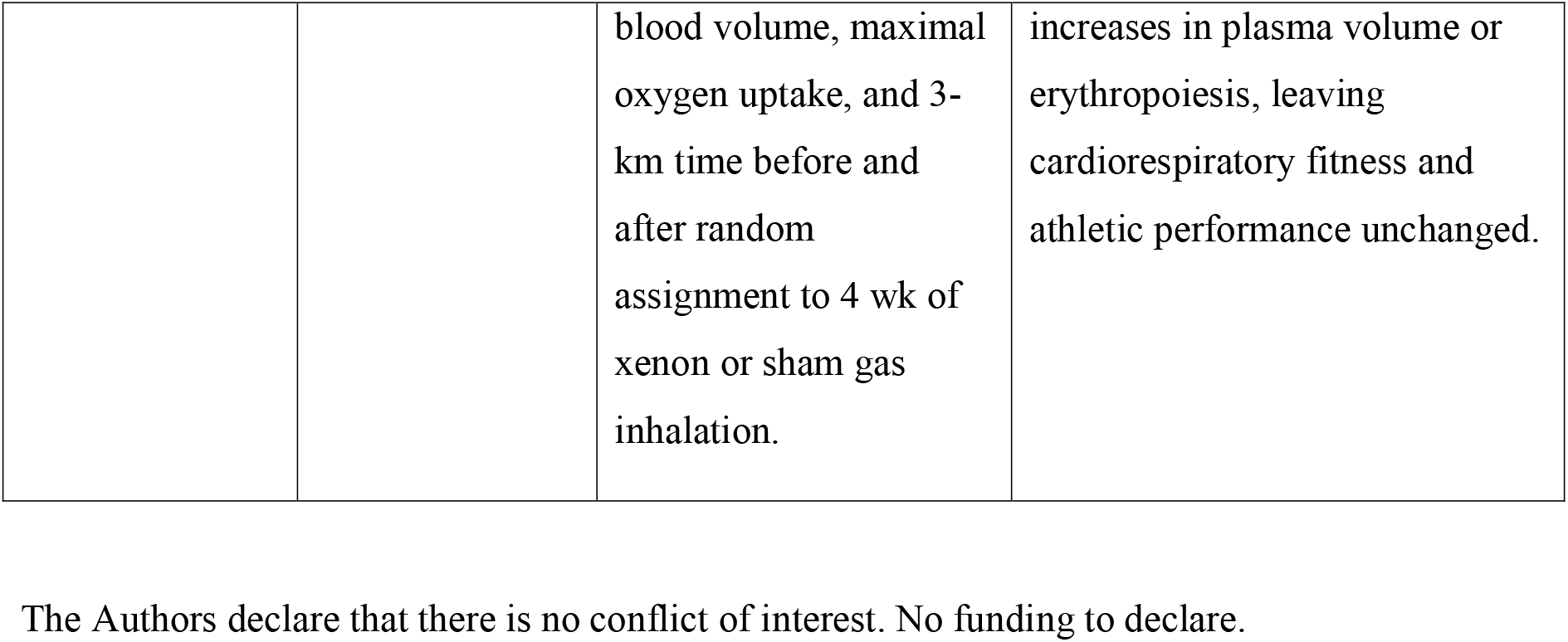
Studies on the effects of xenon and oxygen inhalation on erythropoiesis and steroidogenesis in humans.

| **Article title** | **Number of participants and their physical status** | **Intervention** | **Results** |
| --- | --- | --- | --- |
| Stoppe C et al. 2016 | 24 healthy individuals | Randomly assigned either to a group spontaneously breathing xenon 30 % (Xe/O<sub>2</sub> 30 %/60 %) or a group breathing control gas (N<sub>2</sub>/O<sub>2</sub> 40 %/60 %) for 45 min. | The administration of xenon significantly increased erythropoietin levels 8 h after exposure, peaking at 24 h compared to the baseline values and remained traceable in blood and exhalation probes until 24 h after exposure. In contrast, no significant change was observed in the control group. |
| Dias KA et al. 2019 | 22 healthy individuals | Three subanesthetic concentrations of xenon: 30% fraction of inspired xenon (FiXe) for 20 min, 50% FiXe for 5 min, and 70% FiXe for 2 min. To determine the chronic effects, eight subjects breathed 70% FiXe for 2 min on 7 consecutive days, and EPO, total blood, and plasma volume were measured. Phase II involved assessment of 12 subjects for EPO, total | FiXe 50% and 70% stimulated an increase in EPO at 6 h and at 192 h postinhalation. 7 consecutive days of dosing significantly elevated plasma volume. Phase II showed no significant effect. Acute exposure to subanesthetic doses of xenon caused a consistent increase in EPO, and 7 consecutive days of xenon inhalation significantly expanded plasma volume. However, this physiological response appeared to be transient, and 4 wk of xenon inhalation did not stimulate |
|  |  | blood volume, maximal oxygen uptake, and 3-km time before and after random assignment to 4 wk of xenon or sham gas inhalation. | increases in plasma volume or erythropoiesis, leaving cardiorespiratory fitness and athletic performance unchanged. |

Studies on the effect of IX on steroidogenesis in healthy humans have not been found, nor have studies on the effect of IA on erythropoiesis or steroidogenesis. Studies on the effect of IX on erythropoiesis in mice were published in 2009, 2010, and 2013 and have always been in the public domain. The first study on the effect of IX on erythropoiesis in healthy humans was not published until 2016, and the second study was published in 2019. They involved physically active people from the general population and the number of side-effects reported was minimal (See Table 2).

Prior to 2014, there were only three peer-reviewed scientific studies in English conducted on animals (rats and mice) with simulated pathology or exposed to drugs using long-term high-dose IX inhalation. No studies reporting the effects of IX in humans were published before this date. None of these studies were initiated or funded by WADA. On the WADA website, there is also no study on the effects of IX and IA on erythropoiesis and steroidogenesis.

A search of the largest Russian-language databases did not yield a single publication on the effects of xenon and argon inhalation on erythropoiesis and steroidogenesis. A search for information on athletes disqualified for using or attempting to use IX and IA also failed to find any such cases since 2014.

From the analysis performed, the risk of bias in the included studies is high (See Figure 2). It is important to note that in a study by Dias et al. the randomised trial investigating the effects of xenon was only the phase II of the study (phase I was not randomised). Therefore, we assessed the risk of bias only in the second phase of this study.

**Figure 2.**
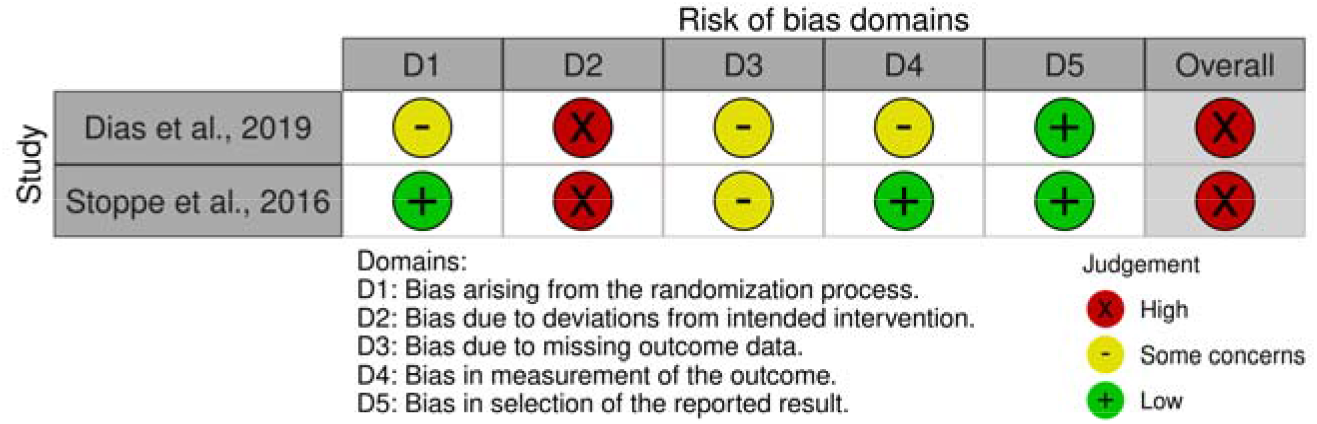
Risk of bias in the studies included.

The main criterion that influenced the result of the risk of bias analysis was D2 (Bias due to deviations from intended intervention). In both studies about 14% of participants were excluded after randomisation and their data were not analysed further. This may have significantly affected the outcome in the context of small sample studies.

## Discussion

A search of English-language scientific databases found only two studies involving healthy human subjects investigating the effects of xenon inhalation on erythropoiesis, both of which were conducted and published after the inclusion of IX and IA on the Prohibited List. They involved physically healthy volunteers and the results either did not support the efficacy of IX as a modulator of erythropoiesis [16] or were obtained using potentially misleading statistical analysis [41],[4],[42]. Furthermore, as the analysis shows, both articles had a high risk of bias.

With regard to the effect of IA on erythropoiesis or steroidogenesis, no studies have been found in the academic domain. Until 2014 there were no studies on the possible effects of IX on erythropoiesis in humans where Jelkmann et al. stated that “effects of xenon treatment on the blood level of EPO have never been reported. No human data are available for the HIF system and the production of EPO” [19]. The author of this review acknowledged that compared to the numerous chemicals that increase HIF-dependent EPO synthesis in humans, the misuse of xenon in sport is a small problem [19].

It is important to note that before the banning of IX and IA in 2014, only three studies on the effects of IX on erythropoiesis were published and both were involving animals (rats and mice) with simulated pathology. Ma et al. demonstrated dramatic effects of xenon inhalation on both EPO levels and levels of HIF-1, a protein that stimulates EPO production in the body. In this study, the authors showed that exposure to a mixture of 70% xenon and 30% oxygen for 2 hours resulted in a sustained increase of HIF-1α activity in adult mouse kidney and human kidney cell line by enhancing the efficiency of HIF-1α translation involving the mTOR pathway [29].

It is important to note that such a protocol of using IX (70% for 2 hours) xenon has never been employed in studies involving humans. Thus, Stoppe et al, stated that inhalation of 30% xenon and 70% oxygen used once a day for 45 minutes [41] was beneficial, and in a study by Dias et al, the maximum concentration of xenon was 70%, and inhalation was performed for 2 minutes [16].

In a study involving male and female mice, Limatola et al. studied the effects of inhaling a mixture of 70% xenon and 30% oxygen for 2 hours on functional neurological outcome and cerebral infarct size after the onset of cerebral ischemia induced by middle cerebral artery occlusion. It was found that both females and males who received IX had a better functional outcome on the focal deficit scale and a smaller cerebral infarct volume. A stronger enhancement of HIF-1alpha compared to the control group who received 70% nitrogen inhalation was also reported [28].

In 2013, Jia et al found that alpha HIF-2 α levels in mice that received 100 mg of the antibiotic gentamicin daily for 7 days remained high for 48 hours after IX application (70% xenon and 30% oxygen for 70 days), while mice placed in a low-oxygen chamber showed increases in EPO that lasted less than 2 hours. However, the authors also found that pre-treatment with IX did not activate hypoxia-inducible factor 1α (HIF-1α) [20].

The first study investigating the effect of IX on erythropoietin levels in humans was published in 2015, one year after it was banned. Stoppe et al. used data from 30 patients from an earlier randomized control trial demonstrating the safety of xenon anesthesia in aortocoronary bypass surgery [39]. The authors noted that although erythropoietin concentrations after xenon anesthesia showed a significant increase on the first post-operative day, this did not result in a statistically significant increase in hemoglobin levels. The authors concluded that the observed association between xenon and erythropoietin and hemoglobin changes remain speculative and causes may be multi-factorial. Stoppe et al. further stated that the results can only be considered as a hypothesis. Interestingly, this was the first and only study that we found examining the possible effect of IX on testosterone levels where no significant changes were found [39].

The first study investigating the effects of IX on erythropoiesis, involving healthy people (24 physically active volunteers), was only published in 2016 - 2 years after the substance was banned. However, the study itself, according to the article, was also conducted post-2014 [41]. In this randomized controlled trial, Stoppe et al. concluded that xenon increased erythropoietin levels in healthy volunteers, based on which the authors considered justification for placing IX on the WADA banned list. The authors also showed increased erythropoietin levels in healthy volunteers after an acute exposure of inhaling xenon for 45 minutes [41].

However, Balachandran et al. commented on these findings and convincingly highlighted that the statistical methods used by the authors do not allow the conclusion that IX positively affects erythropoiesis in humans to be drawn. The main problem with the statistical analysis pointed out by Balachandran et al. was that the conclusion about the effectiveness of xenon was based on comparing the significant within-group change observed in the xenon group with a small change in the control group, and no comparison was made between groups [4]. However, the practice of comparing p-values within a group can be misleading [9]. The authors of this commentary emphasized that if a trial is to claim superiority, statistical tests comparing average differences between groups, i.e. group analysis of variance or two-sample t-test, should be performed and the conclusion that xenon increases EPO levels in humans is inappropriate and possibly misleading [4]. In their response, Stoppe et al. acknowledged that “the observed results should be investigated more thoroughly in subsequent confirmatory studies” [42].

In 2019, 5 years after IX was banned; a study by Dias et al. was published that examined the acute and chronic effects of various IXs on 12 physically active volunteers from the general population, not athletes (which the authors themselves considered to be a significant limitation). The authors of the study particularly noted that it was “the first study to examine each element of the cascade by which xenon inhalation is purported to take effect, starting with measurement of the hypoxia-inducible factor effector, erythropoietin, to hemoglobin mass and blood volume and athletic performance” [16]. In this study, three different IX protocols were used: a 30% fraction of inspired xenon for 20 minutes, 50% for 5 minutes, and 70% for 2 minutes. The results showed that IX at 50% and 70% fraction of inspired xenon increased the EPO concentration after 6 hours and 192 hours after a single application. However, application of IX at a concentration of 70% for 7 days showed no significant effect on EPO, hemoglobin mass, plasma volume, maximal oxygen uptake, or a 3-km time-trial. The authors concluded that the physiological response of IX was temporary and 4 weeks of xenon inhalation did not stimulate increases in plasma volume or erythropoiesis, leaving cardiorespiratory fitness and athletic performance unchanged. Authors also found that acute xenon inhalation caused small prolonged increases in EPO, but short-term daily exposure did not provide superior benefits beyond an acute dose. The increase in plasma volume with short-term daily dosage was found. Authors concluded that xenon inhalation did not improve athletic performance, and stated that their findings do not support the use of xenon as an erythropoiesis-modulating agent in sports [16].

It should be noted that the protocols used in studies involving healthy individuals differed significantly from those described in the Economist article in 2014. It is also interesting to note that Dias et al. used the opinions of some Russian trainers to select the protocols for IX use, rather than the protocols previously described in studies involving humans and animals. That is, at the time of the study there were no scientifically valid protocols for the use of IX in healthy volunteers to realize a potentially positive effect on performance and erythropoietin levels [16].

In the Dias et al. study, a statistically significant increase in plasma volume was shown when using IX at a concentration of 70% for 2 minutes on 7 consecutive days. However, an increase in plasma volume alone does not cause a positive effect on performance, as evidenced by the lack of prohibition of other methods actively used by athletes that increase plasma volume. For example, Russian endurance athletes actively use sauna [7], which, according to Scoon et al. can potentially increase plasma volume by more than 7%. It will be interesting to note that in the same study, the authors showed that a 30-minute visit to a sauna for 3 weeks increased the ability to run to exhaustion by 32%, which may be equivalent to a 1.9% improvement in competitive running [37]. However, sauna remains a legal recovery method and is not included on the Prohibited List.

It is also important to remember that an improvement in endurance performance and a significant increase in hemoglobin (of approximately 6%) with recombinant erythropoietin, which has long been strictly prohibited in sport, is only observed 2 weeks after the administration of recombinant EPO [37]. Therefore, an estimate of low and short-term erythropoietin levels with IX use cannot be considered as a reliable effect on physical performance, as was shown in the Dias et al. study in which the authors saw no positive effect of IX on performance in the 3-km run [16].

A search of the largest Russian-language databases also failed to find any articles on the effects of IX and IA on erythropoiesis and steroidogenesis. In addition, it should be noted that no significant adverse health effects have been observed in studies involving the use of xenon in humans, and its main global use is as an anesthetic in surgical procedures, including cardiac surgery. The possible non-compliance with the spirit of the sport of these particular inhalations is also questionable, given that helium and oxygen inhalation, for example, remain permitted and are actively used in Russia, including among athletes, but are not on the WADA Prohibited List [5],[6].

Another possible explanation for the inclusion of IX and IA on the Prohibited List could be the widespread use of IX and IA by a particular group of athletes. To objectively determine the prevalence of a substance or method, accurate detection methods must be used. In July 2014, a study led by Thevis et al., was published showing that xenon can be determined from plasma and blood samples, i.e. common samples of routine sports drug testing with detection limits of approximately 0.5 nmol/mL to 50 nmol/mL, depending on the type of mass spectrometer used [5]. This article showed that the detection of IX is possible using standard detection methods and provided data on the detection limit of xenon.

Furthermore, Thevis et al. presented the first data on the determination of xenon from urine in the context of human sports drug testing [46]. This publication also stated that, given inclusion on the Prohibited List, testing for xenon in samples submitted for doping control is required and presents the first data on the determination of xenon in human urine during testing for sports drugs, where indicating a detection limit of approximately 0.5 nmol/mL and a detection time of approximately 40 hours after xenon anesthesia [46]. In Schaefer et al, study the xenon detection time after standard anesthesia using an IX concentration of 60% was 24 -48 hours [35]. Thus, it is questionable that during the 2014 Sochi Olympics sports organization obtained accurate information utilizing these methods.

Apparently, IX and IA have never been actively used and are not used by athletes to enhance erythropoiesis, because in the 8 years since their prohibition there have been no disqualifications for their use, while in recent years there are accurate and objective methods for their detection in bodily fluids. Simultaneously, disqualifications for the use of other agents and methods that actually enhance erythropoiesis (e.g. EPO) have been extremely frequent.

In summary, there were no data to suggest the real prevalence of IX in athletes of any nationality, or any data on the effects of IX on erythropoiesis and steroidogenesis in humans. The only data available on the effects of IX on erythropoiesis is based on studies in mice and rats with simulated pathology. Therefore, it is unclear why these substances and methods were included on the Prohibited List.

After a structured re-evaluation of the 2020 Prohibited List, argon was removed “because it is considered to no longer meet the criteria for inclusion” [50]. While in 2022 the Prohibited List maintained xenon as a prohibited HIF-activating agent [51]. Xenon remains prohibited, despite the fact that over the past 8 years there has been no evidence of its positive effect on erythropoiesis and steroidogenesis, as well as data suggesting its wide prevalence prior to its inclusion on the prohibited list. There is also no evidence to support the notion that IX has a negative health impact and there is not a single case of dis-qualification for its consumption, which raises the question of its significance in enhancing any aspect of performance.

We believe that publishing data that justifies the decision to include the substance or method to the Prohibited List will promote the trust of the stakeholders. Showing evidence behind these decisions would promote process transparency and reduce the opportunity for speculation [8]. The possibility of developing clear guidelines (e.g. the performance of RCTs and research in detection methods in bodily fluids) prior to the inclusion of various substances and methods can be considered.

## Conclusion

Based on existing literature, there is no evidence that xenon inhalation can increase testosterone levels, and studies that have examined the effect of xenon inhalation on erythropoietin levels are inconclusive. In all the studies conducted there is no evidence of any significant adverse health effects. For IA, there is still no evidence to suggest it has any positive effects on steroidogenesis and erythropoiesis or any adverse health effects. Therefore, WADA should either enable high-quality studies to provide reliable and unambiguous data on the effects of IX on erythropoiesis and steroidogenesis or remove this substance from the Prohibited List. Finally, a study with retrospective analysis of all doping samples of Russian competitors at the Sochi Olympic Games for xenon should also be undertaken to determine the actual prevalence of IX use at these events.

## Data Availability

All data produced in the present study are available upon reasonable request to the authors

## Notes

### Competing Interest Statement

The authors have declared no competing interest.

### Funding Statement

This study did not receive any funding

### Summary of Updates

Methods and Results sections (more databases and libraries for research, some irrelevant information to remove)

